# Early recovery of leukocyte subsets is associated with progression-free survival in patients with inoperable stage III NSCLC after multimodal treatment: a prospective explorative study

**DOI:** 10.1101/2023.11.16.23298143

**Authors:** Thomas P. Hofer, Alexander E. Nieto, Lukas Käsmann, Carolyn J. Pelikan, Julian Taugner, Saloni Mathur, Chukwuka Eze, Claus Belka, Farkhad Manapov, Elfriede Nößner

## Abstract

**Background:** We explored the dynamic changes of major leukocyte subsets during definitive treatment of patients with inoperable stage III NSCLC lung cancer and correlated it to survival to identify subpopulations associated with maximal patient benefit.

**Methods:** We analyzed peripheral blood of 20 patients, either treated with thoracic radiotherapy (RT), concurrent chemo-radiotherapy (cCRT), or cCRT with additional immune-checkpoint inhibition therapy. Blood samples were collected at 9 timepoints before, during, and up to 1 year post treatment and analyzed by multi-color flow cytometry. Statistical analysis was conducted for leukocyte subpopulations, IL-6, progression-free survival (PFS) and overall survival (OS).

**Results:** Increase of absolute lymphocyte counts (ALC) after the end of RT until 6 months thereafter was a predictor of PFS. Baseline lymphocyte counts showed no significant correlation to PFS or OS. Early recovery of absolute counts (AC) at 3 weeks after RT, total CD3+ T-cells, and CD8+ cytotoxic T-cells distinguished those patients with favorable PFS (≥12 months) from all other patients. Discriminant analysis identified B-cells, neutrophil-lymphocyte-ratio (NLR), CD4+ T-helper-cells, and NK-cells as predictors of favorable PFS. High variability in IL-6 plasma concentration of consecutive measurements within 6 months after the end of RT correlated negatively with PFS.

**Conclusion:** Our results suggest that two parameters commonly assessed in clinical routine, can be used to predict patient outcome. These are: early increase in CD8+ T-cell lymphocyte-count and variability in IL-6 plasma concentration, that are correlated to patients with favorable, respectively, poor outcome after definitive therapy independent of treatment regimen.

**Highlights:**

- Early increase within 3 weeks after thoracic radiation therapy (TRT) of CD8+ T cells is associated with favorable progression free survival (PFS).
- Low standard deviation in IL-6 plasma concentration in consecutive measurements after TRT is associated with favorable PFS.
- Absolute lymphocyte counts at treatment begin had no predictive value for treatment outcome.

**Funding statement:** none

## Background

Non-small cell lung cancer (NSCLC) accounts for approximately 85% of all lung cancers, and prognosis remains poor with a low 5-year survival rate, only up to 36% of patients with an advanced stage experiencing sustained clinical benefit from therapy [1]. Treatment advances were achieved by introducing immune checkpoint therapy (ICI) as monotherapy or in addition to chemo-radiotherapy [2, 3].

There is a medical need to not only find more effective therapies but also to identify those patients who are likely to benefit from added ICI or should be spared the side effects if ICI is ineffective [4]. Ongoing efforts are being made to identify suitable biomarkers to predict benefit or non-response, preferably obtained from easily and repeatedly accessible biomaterial like venous peripheral blood.

In this prospective single-center study we aimed to explore whether the dynamics of defined lymphocyte subsets might be suitable to predict survival. We performed immune monitoring on patients who underwent therapy for inoperable stage III NSCLC. We prospectively report on characteristics of absolute T-cell counts (total CD3+ T-cells, CD3+CD4+, and CD3+CD8+ T-cells), B- cells, NK-cells, neutrophils, thrombocytes, and IL-6 plasma levels regarding patient treatment and outcome (progression free survival, PFS, and overall survival, OS).

## Material and Methods

### Patients, Controls, Blood Sampling, Treatment

Blood samples from patients with unresectable stage III non-small cell lung cancer (NSCLC; 8^th^ TNM staging system) without distant metastases were obtained before as well as during treatment and follow-up at the Clinic for Radiation Oncology of the LMU University Hospital, Munich. All patients were included in this study on a voluntary basis and provided written informed consent. The study was approved by the Human Ethics Committee of the Ludwig-Maximilians-University of Munich reference no. 17-632 and conducted in accordance with the Declaration of Helsinki.

Twenty patients (17 male, 3 female), first diagnosed with a median age of 65.5 years (range 34 to 79) were enrolled in this study (Tab. 1; Fig. S1).

**Tab. 1:** Characteristics of patients with NSCLC and healthy controls.

| Patients | n=20 |
| --- | --- |
| Age (mean SD) (years) | 62.5 ± 13.75 |
| (median / range) (years) | 65.5 / 34-79 |
| Sex (male / female) | 17 / 3 |
| Therapy (RT, cCRT, cCRT-ICI, salvage therapy) | 2 / 12 / 7 / 3 |
| Adenocarcinoma | 11 |
| Squamous cell carcinoma | 8 |
| Undiff. NSCLC | 1 |
| Distant metastasis | 8 |
| Local remission | 9 |
| Death | 7 |
| Smokers (none / active / ex) | 1 / 7 / 12 |
| Smoking history > 50 pack years | 8 |
| Healthy Controls | n= 9 |
| Age (mean SD) (years) | 35 ± 13.74 |
| (median / range) (years) | 30 / 24-59 |
| Sex (male / female) | 1 / 8 |
| Smokers (none / active / ex) | 8 / 1 / 0 |

Blood samples were collected before onset of therapy (A.1 at week 0), twice during (A.2 and A.3 at week 2 and 5, respectively) and at the end of radiotherapy (RTend at week 7), and during follow up (C.1 to C.5 at week 10, 20, 35, 48, and 60) (Fig. S1).

The controls constituted of 9 apparently healthy individuals (CTRL) with a median age of 30y (range 24 to 59) at the timepoints of blood sampling; 1 male and 8 females (Fig. 1, Tab. 1). Samples were processed and analyzed simultaneously and identically to the patient samples.

**Fig. 1.**
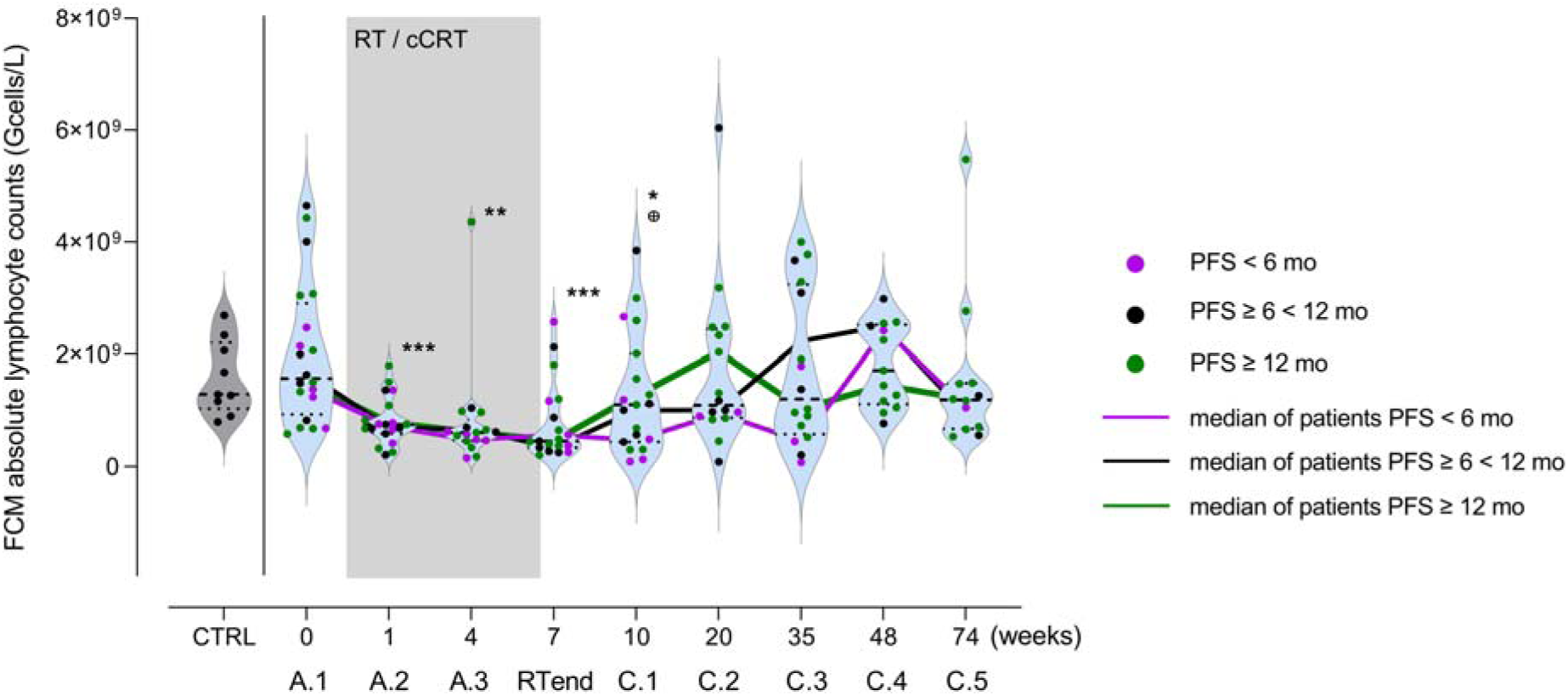
ALC in NSCLC patients’ peripheral blood (n=20) before (A.1), during (A.2, A.3, RTend), and up to 1 y (C.1-C.5) after treatment. Grey violin represents ALC of CRTL (n=9). Each dot represents an individual, color codes the PFS group of the patients. Grey area represents the interval of radio-(RT) or chemoradiotherapy (cCRT), color-coded lines connect the median of ALC of each PFS group at the different timepoints. The bold dashed line in the violins indicate median, light dotted lines the lower and upper quartile. Student’s t test, * p<0.05, ** p<0.01, ***p<0.001 compared to A.1; ⊕ p<0.05 compared to preceding timepoint.

Patient treatment modality was concluded at the discretion of the treating physician based on clinical standards and following multidisciplinary tumor board discussion. Two patients received RT alone, 18 patients were treated with platinum-based concurrent cCRT, and 7 patients received additional ICI either concurrently (nivolumab) or sequentially (durvalumab). More detail is provided in Fig. S1.

### Leukocyte Phenotyping

Blood was collected using 3K-EDTA S-Monovette tubes (Sarstedt, Germany) and samples were analyzed within 3 hours. Using 100 µl of whole blood, erythrocytes were lysed, cells were surface-stained, spiked with counting beads and analyzed on a BD LSRII flow cytometer. Details on sample preparation, staining and gating strategy, cell type identification are given in the supplements (Fig. S2, Tab. S1).

Neutrophil and lymphocyte counts used to calculate Neutrophil-Lymphocyte-Ratio (NLR), eosinophil counts, and IL-6 plasma levels were obtained from clinical routine blood testing (clinical chemistry).

### Analysis of immunocyte population dynamics and variation of IL-6 plasma concentration

To describe the dynamic changes in absolute cell counts during the period after treatment end (RTend), we calculated the area under the curve (AUC) of consecutive blood sampling timepoints from RTend-C.3 (6 mo after treatment end). Low or high AUC reflect higher or lower AC in this period.

For analysis of variation of IL-6 concentration during RTend-C.3, standard deviation of absolute values at three consecutive timepoints after RTend were calculated and stratified according to groups with poor (<6 mo), intermediate (≥6<12 mo), and favorable (≥12 mo) PFS.

### Statistical analysis

This study was designed as a discovery study with the intention of generating hypotheses. Statistical tests were Student’s t-test, Kruskal Wallis test, Log-rank (Mantel-Cox), Fisheŕs Exact test, and Log-Rank for trend. Differences were considered significant with p<0.05. For multivariate discriminant analysis (MDA), and heatmap visualization, R statistical software V4.2.1 and the packages ComplexHeatmap V2.12.1, Dplyr V1.0.10, Plyr V1.8.7, missMDA V1.18, Openxlsx V4.2.5, Stats V4.3.0 and Tidyverse 1.3.2 were used.

For MDA, we used the ggord package from Marcus W. Beck (2017). ggord: Ordination Plots with ggplot2. R package version 1.0.0 https://zenodo.org/badge/latestdoi/35334615. For generating graphs and Kaplan-Meier curves, GraphPad Prism V9, and for MANOVA analysis Past V4.11 [5] were used.

## Results

### No prognostic value of ALC for PFS or OS at any single timepoint before, during and after treatment

Baseline ALC were highly heterogenous in the NSCLC patient cohort (n=20) with a higher median of 1.57 (range 4.65-0.58) Gcells/L of ALC compared to the controls (n=9, median 1.29, range 2.69-0.80 Gcells/L). All patients experienced moderate to severe lymphocytopenia (assessed per the Common Terminology Criteria for Adverse Events v5.0) during RT/cCRT with median ALC of 0.75 (range 1.80- 0.21), 0.61 (range 4.36-0.15), and 0.46 (range 2.57-0.21) Gcells/L at timepoint A.2, A.3, and RTend, respectively (grey area in Fig.1). ALC recovered after RTend reaching median values of 1.10 (range 3.85-0.09) Gcells/L at C.1 (3 wks after RTend), 1.09 (range 6.04-0.09) Gcells/L at C.2 (3 mo after RTend), 1.21 (range 4.00-0.08) Gcells/L at C.3 (6 mo after RTend), 1.72 (range 2.99-0.77) Gcells/L at C.4 (10 mo after RTend), and 1.19 (range 5.47-0.54) Gcells/L at C.5 (13 mo after RTend). The recovery varied across patients, some with poor others with good increase in cell counts.

Patients were stratified according to PFS after RTend, with poor PFS<6 mo indicated by purple dots, intermediate PFS≥6<12 mo shown as black dots, and favorable PFS≥12 mo depicted as green dots.

Patients with favorable PFS had on average an early increase in ALC within 3 weeks after RTend (C.1), peaking at timepoint C.2 (14 weeks after RTend) and return to a median ALC comparable to the CTRL. Patients with poor and intermediate PFS had a delayed increase in ALC with a peak in cell counts at C.4 (40 weeks). Although these data indicated that an early recovery of ALC may be beneficial with regards to PFS, there was high ALC heterogeneity within the PFS groups: 3 of 6 patients in the favorable PFS group had no early ALC peak at 14 weeks.

### Dynamics of leukocyte subpopulations after RT

Favorable PFS group (≥12 mo) showed significant increase in AC from RTend to C.1 for lymphocytes, total T-cells, and CD8+ T-cells. For the other PFS groups, lymphocytes and CD4+ T-cells did not increase, or the increase occurred later (between RTend and C.3) (Fig. 2, Fig. S4). This was observed for total lymphocytes as well as total CD3+ T-cells and CD8+ T-cells, but not for CD4+ T-cells (for medians and p-values see Tab S1). This implies that an early increase of lymphocytes, total CD3+ T-cells, or CD8+ T-cells within 3 or 14 weeks after RTend (C.1, or C.2) identifies patients with a favorable progression-free survival-time ≥12 mo.

**Fig. 2.**
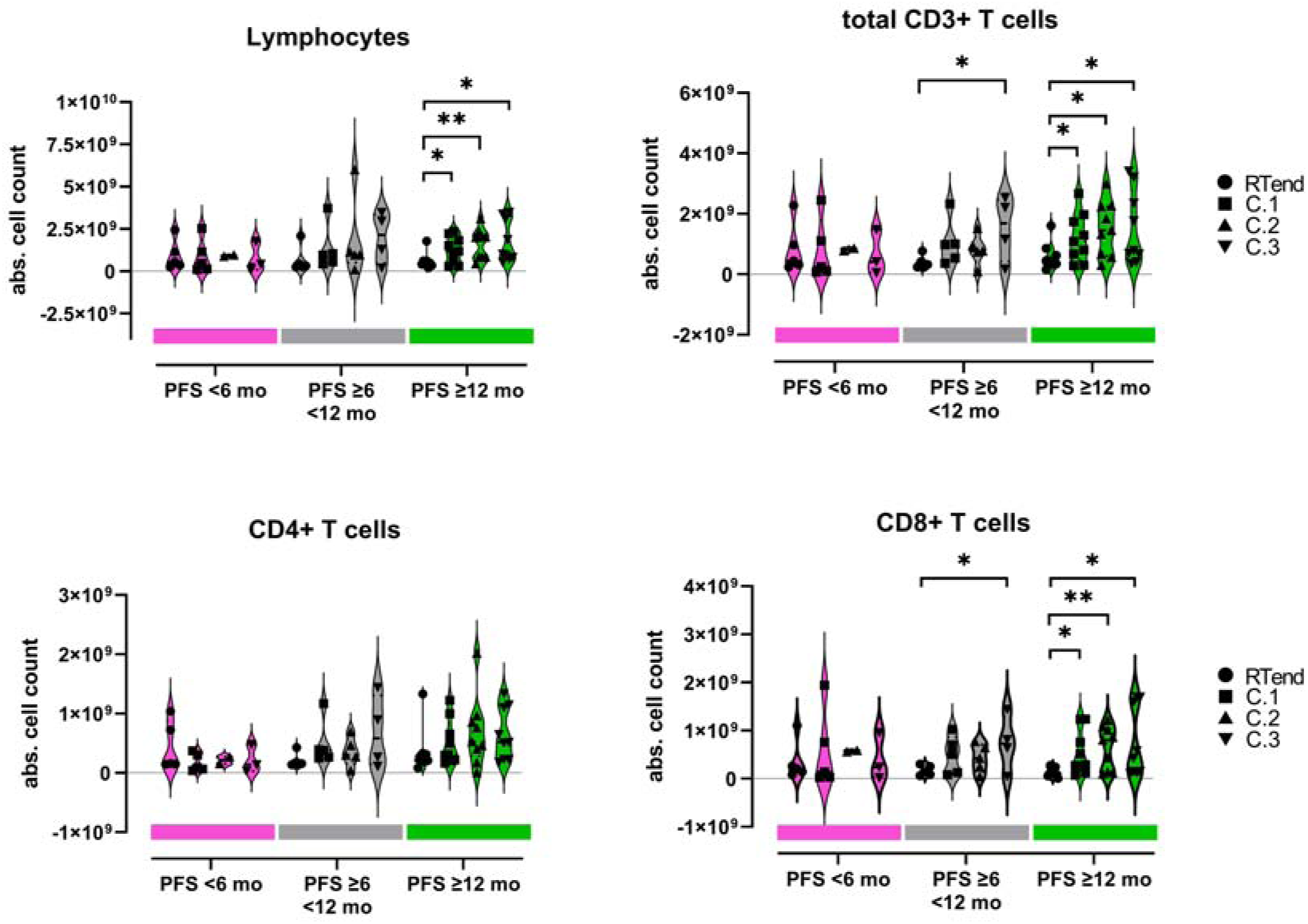
Absolute cell counts of lymphocytes, CD3+ T-cells, CD4+ T-cells, and CD8+ T-cells (analyzed with flow cytometry) at timepoints RTend, C.1, C.2, and C.3 within the different PFS groups (PFS<6 mo (red), PFS ≥6<12 mo (grey), PFS ≥12 mo (green)). Cell counts were derived from flow cytometry (gating strategy in Fig. S2), each dot represents an individual. Comparison was performed across timepoints within each PFS groups, bold dashed line in the violins indicate median, light dotted lines lower and upper quartile. Student’s t-test, * p<0.05, ** p<0.01.

An increase in neutrophil count with inadequate T-cell and no NK-cell expansion after RTend was observed in the poor prognostic group (Fig. 2, Fig. S4). The intermediate and favorable prognostic groups showed peripheral NK-cell increase after RTend (Fig. S4) while the poor prognostic group did not (median value for intermediate group at RTend 0.014, at C.1 0.040, at C.2 0.132, at C.3 0.257 Gcells/L; RTend-C.1 p=0.094, RTend-C.2 p=0.011, RTend-C.3 p=0.015; median value for favorable group see Tab. S2). The favorable prognostic group showed increaseF after RTend of CD8+ T-cells (median value for intermediate group at RTend 0.14, at C.1 0.51, at C.2 0.42, at C.3 0.73 Gcells/L; RTend-C.1 p=0.0596, RTend-C.2 p=0.053, RTend-C.3 p=0.031; median value for favorable group Tab. S2).

All patients experienced leukocytopenia following RT or cCRT (Fig. 1). We hypothesized that the change of lymphocytes within the first 6 mo after end of therapy might be most meaningful for therapy response as the immune system reconstitutes and can react to neoantigens released from destroyed tumor cells. To picture the dynamic processes during this phase between RTend-C.3 (6 mo), AUC of the absolute cell counts of the major leukocyte subpopulations was calculated and compared to patient outcome.

Higher AUC for the period between RTend-C.3 reflect a more pronounced increase of absolute cell counts after therapy. Patients in the poor PFS group (PFS<6 mo) compared to the favorable prognostic group (PFS≥12 mo) had significantly lower AUC for CD3+ T-cells (p=0.0053), CD4+ T-cells (p=0.0026), and NK-cells (p=0.0008) (Fig. 3). AUC of CD8+ T-cells, B-cells, eosinophils, and NLR were not different between the poor and good prognostic groups.

**Fig. 3.**
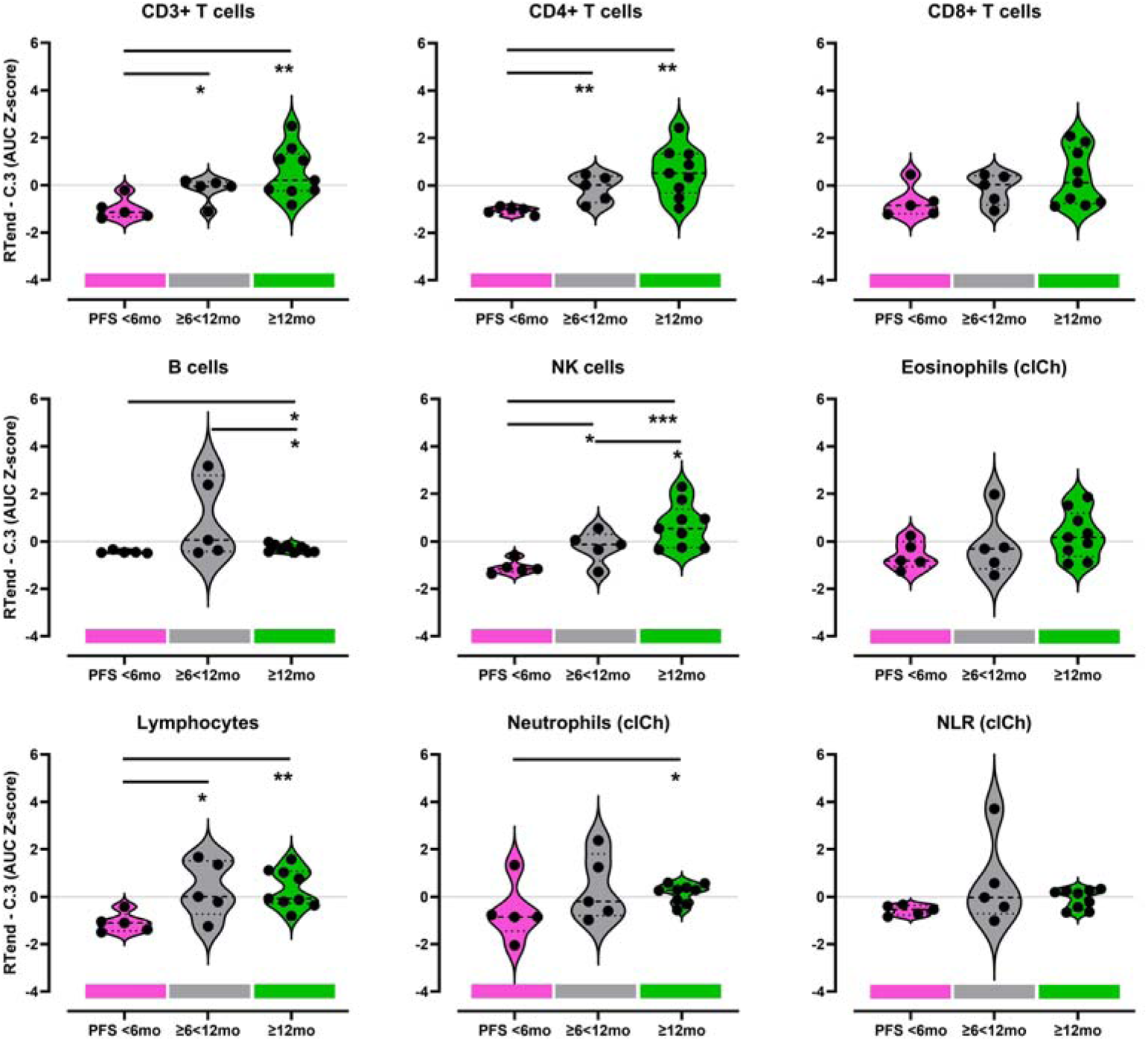
Standardized AUC (Z-score) for the interval RTend-C.3 of leukocyte subpopulations stratified according PFS groups (PFS<6 mo (red), PFS ≥6<12 mo (grey), PFS ≥12 mo (green)). Cell counts were derived from flow cytometry (gating strategy in Fig. S2), neutrophil, and eosinophil counts were determined by clinical chemistry routine lab (clCh). Each dot represents the value of an individual, bold dashed line in the violins indicate median, light dotted lines lower and upper quartile. Student’s T test, * p<0.05, ** p<0.01, ***<0.001.

### Unsupervised clustering reveals longer PFS for patients receiving ICI

Normalized lymphocyte parameters were clustered by unsupervised hierarchical clustering. PFS, treatment regimen, local relapse, distant metastases were annotated to, but were not included in the clustering. The result (Fig. 4) identified two clusters: on the right side were patients with low AUC for all cell types, except NLR (assigned “cold” group) (9 of 19 patients). Only 1 patient (1 of 9 patients) within this group had received ICI. Most patients (7 of 9patients) in this “cold” lymphocyte-reduced group had developed local tumor recurrence (LR) after definitive therapy and only 2 (of 9) belonged to the favorable group (PFS≥12 mo). Patients clustering to the left (10 of 19 patients) had higher AUC (RTend-C.3) for all leukocyte subpopulations except NLR (assigned “hot” group). Six of the patients of the “hot” group had been treated with ICI (6 of 10), only 2 (of 10) had experienced LR and most belonged to the favorable PFS group (7 of 19).

**Fig. 4.**
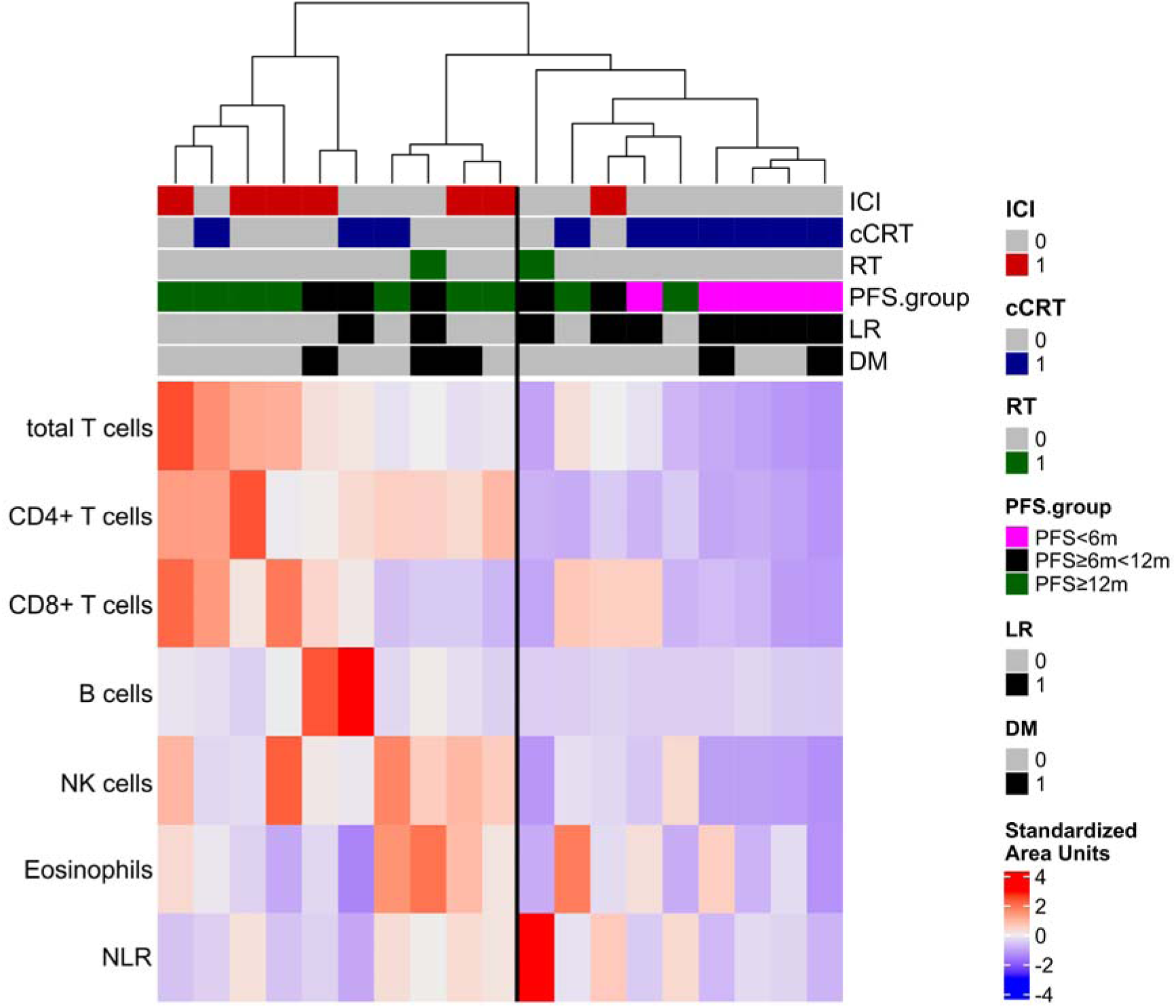
Heatmap of unsupervised clustering of AUC (RTend-C.3) of leukocyte subpopulations, every column represents a patient, PFS (<6mo, ≥6 <12mo, and ≥12mo), treatment regime and recurrence (LR, local recurrence; DM, distant metastases) (AUCs) in patients (n=19). Bold vertical line separates a “hot” group (higher AUC) and a left “cold” group (lower AUC).

Fisher’s Exact Testing “cold” or “hot” group against good or poor PFS (≥12 mo 10 patients; and <12 mo, 9 patients, respectively) revealed a statistically non-significant value of p=0.069. The Odds ratio (95% CI) was 7.17 (0.752, 113.47).

### Linear discriminant analysis (LDA) using blood cell subpopulations separates patients according to PFS

We analyzed the AUC of cell counts (CD4+ and CD8+ T-cells, B-cells, CD56+CD3- NK-cells, eosinophils, neutrophils, and the NLR) at the RTend to C.3 interval as predictor variables for PFS (response variable), defined in 3 groups: PFS<6 mo, ≥6<12 mo, and ≥12 mo.

Discriminant markers for PFS≥12 mo (green area) were the AUCs of CD4+ T-cells, and CD56+ NK-cells (Fig. 5, Fig. S5). By these markers, 6 of 9 patients of the PFS≥12 mo group were correctly separated from the other PFS groups. Patients with intermediate PFS (≥6<12 mo) (grey area) were discriminated by AUCs of B-cells and NLR, with four of five individuals of this group separated correctly. The mean of prediction as a measure for accuracy was 0.947 for linear discriminant analysis.

**Fig. 5.**
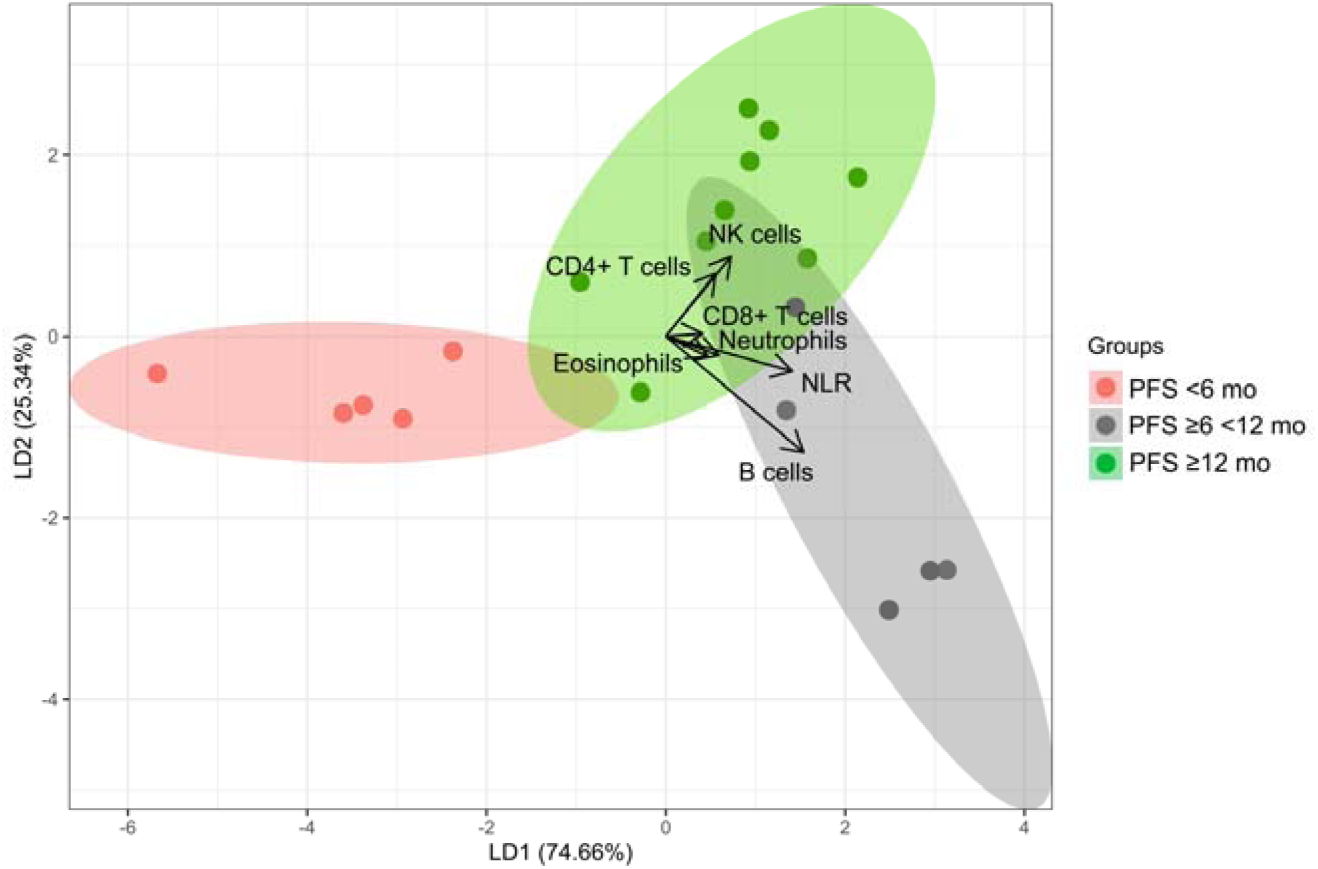
Linear discriminant analysis group plot (response variable PFS, predictor variables were AUCs of the interval RTend-C.3 of CD4+ T-cells, CD8+ T-cells, B-cells, NK-cells, eosinophils, neutrophils, and NLR). Mean as a measure for accuracy was 0.947. The length and direction of the arrows indicate how well the parameter differentiates this group from the others.

**Fig. 6.**
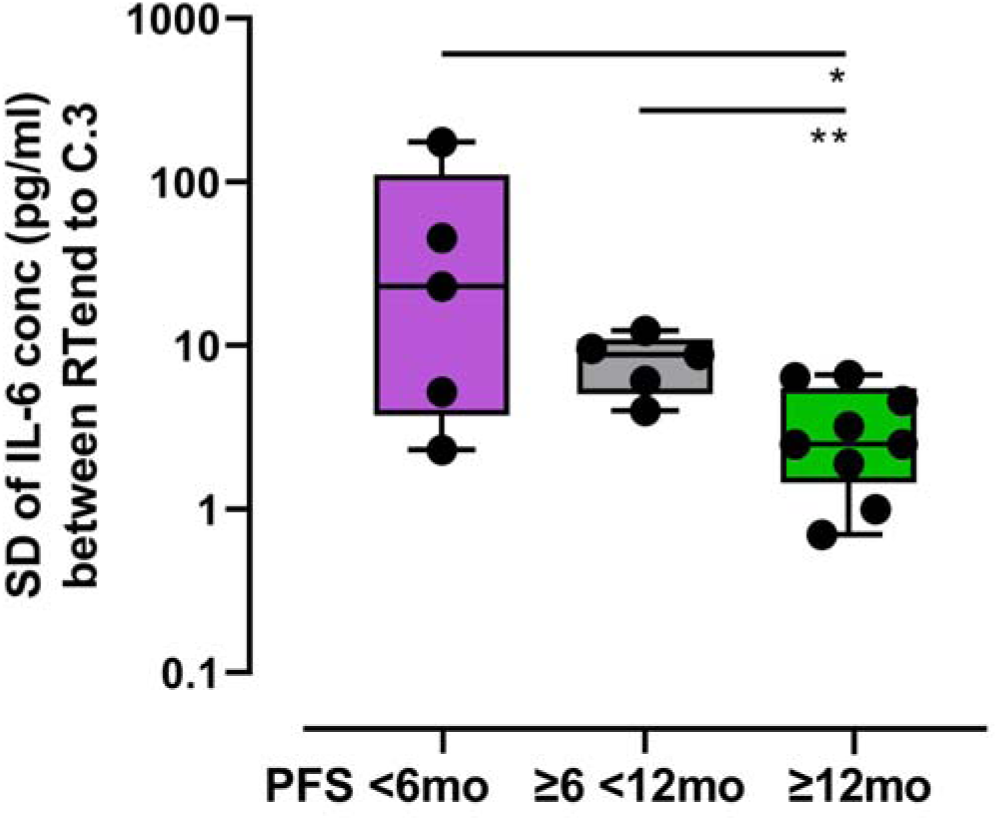
Standard deviation (SD) of IL-6 concentration from RTend-C.3 according to PFS groups (box plots showing median, min and max); Student’s T test, * p<0.05, ** p<0.01.

### Variation in IL-6 concentration is correlated with PFS

IL-6 is a pro-inflammatory cytokine that plays an important role in innate immunity and is upregulated in sepsis [6], after trauma [7], and medical treatment, e.g. surgery [8]. Basal IL-6 levels highly vary inter-individually [9], thus a comparison of absolute values for IL-6 across individuals is difficult to interpret. We focused on the intra-individual variation of IL-6 concentrations (pg/ml) over at least 3 consecutive timepoints, starting at RTend until C.3 as a clinically sensible interval for survival prediction. The standard deviation of the IL-6 concentrations (pg/ml) during this interval (n=4 blood samples each patient) was used as a measure of variation of the IL-6 level.

The favorable survival group had a median standard deviation (RTend-C.3) of 2.5 (range 0.7-6.6), the poor survival group 22.9 (range 2.3-175.7), and the intermediate group 8.8 (range 4.0-12.3). Thus, the IL-6 variation after RTend was significantly lower for the favorable PFS group and this significantly correlated with PFS (p=0.007, Fig. 6).

## Discussion

In this prospective explorative study, we evaluated the dynamics of defined peripheral leukocyte subsets in patients with inoperable stage III NSCLC during primary treatment and follow-up with the aim to identify peripheral T-cell subsets that may predict treatment outcome, especially PFS.

Our analysis provides evidence that ALC expansion after the end of definitive RT or cCRT until 6 mo afterwards is significantly associated with treatment efficacy. Increase of AC in CD3+ T-cells, CD8+ cytotoxic T-cells, and NK-cells had the most pronounced effect and was associated with favorable PFS, whereas significant increase in B-cell counts had a less apparent positive association with PFS and eosinophils and CD4+ T-cell counts had no association.

ALC before treatment (A.1) or at any other single timepoint did not reveal a correlation with PFS (Fig. 1). There was no significant correlation between ALC at six months after RTend and treatment regimen (data not shown).

Comparable to findings of previously published studies [10], [11], [12], [13], all patients in this study experienced moderate to severe lymphocytopenia during RT (A.2-RTend) with the nadir at the end of radiotherapy. Most of our patients recovered from lymphocytopenia within the following 3 (C.2) to 6 (C.3) mo. Some (4 of 16 at C.3) had persistently low cell counts (Fig. 1), which correlated with poor disease control with PFS less than 6 mo (Fig. 3). An earlier study [14] found an association between severe lymphocytopenia (<500 cells/µl) and reduced OS (HR 1.70, p=0.17). Similar evidence was observed in glioblastoma [15], cervical cancer [16] and in seminoma [17]. This is comparable to what we see in this study, where patients with continuously low ALC had poor PFS and OS (Fig. S3c,d).

In our study patients had favorable PFS and OS when treated with cCRT+ICI compared to cCRT alone (Fig. S3e,f). In a comparable investigation, Jing et al. [18] compared NSCLC patients who received cCRT with those who received additionally maintenance treatment with durvalumab. They found a significant increase in PFS in the durvalumab subgroup alongside with a significant correlation between lymphocytopenia and PFS/OS. Jing et al. observed no statistically significant evidence that patients with lymphocytopenia benefited from durvalumab in terms of improved overall survival. However, the authors focused on severe lymphocytopenia with a cell count of less than 0.23 Gcells/L (230 cells/µl). Additional factors may play a role when ALC is extremely low. Our cohort was small and only five patients had very low ALC at the end of radiation. Of these patients, two received RT, one cCRT, and two received cCRT+ICI. Two cCRT+ICI patients showed a strong recovery of cell counts together with favorable PFS (≥23 mo). Interestingly, Cho et al. [12] postulated that patients with severe lymphocytopenia may benefit most from additional ICI because a fresh and immunological potent lymphocyte population may arise from a depleted lymphocyte reservoir.

The timeperiod following end of RT is considered to be significant since processes involving immunogenicity take place that play a role in durable immune activation and tumor rejection [19], [20]. In this study, we focused on the 6-mo period of RTend-C.3 and refer to it as a crucial recovery phase. Considering the different lymphocyte subtypes (Fig. 2), we identified that an early increase in absolute counts after RT of total CD3+ T-cells and CD8+ T-cells was predictive of favorable PFS. Interestingly, Belka et al. [17] found that CD8+ T-cells recovered significantly faster than CD4+ T-cells after end of radiation treatment in patients with seminoma. In head and neck cancer, a gradual increase in CD8+ T-cells over 6 mo after radiotherapy was observed for non-recurrent patients [21]. This supports our finding that an early increase in CD8+ T-cells in peripheral blood may be useful to predict favorable PFS. Kim et al. [22] reported on increasing frequency of Ki67+ cells within CD8+ PD- 1+ T-cells in patients with locally advanced NSCLC during cCRT peaking at the last week during therapy followed by a decline 1-mo post-cCRT. These cells acquired a senescence phenotype. Although we did not include the Ki67 marker in this study we did not see a decline of CD8+ T-cells but rather more CD8+ T-cells in absolute counts from C.1.

We assumed that the dynamic changes of the cell counts may be meaningful for patient survival. Evidence that T-cell recovery is important was shown in a study of melanoma patients by Huang et al. [20]. They found that a CD8+ Ki67+ T-cell subpopulation positively correlated with clinical outcome when it peaked at a given early timepoint after therapy begin and subsequently decreased. To reflect the dynamic changes in absolute cell count in given subpopulations during the recovery interval of Rtend-C.3, we calculate the AUC between four consecutive timepoints within 6 mo after RT (RTend-C.1-C.2-C.3). This new approach can depict a broader picture of cellular dynamics in peripheral blood than the stepwise comparison of single timepoints. We identified that total lymphocytes and neutrophils, CD3+ T-cells, B-cells and NK-cells recover significantly faster and reach significantly higher AUC in patients with favorable PFS≥12 mo. Considering the T-cell subsets in this study, the prediction of long-term survival using this AUC method was statistically significant with the CD4+ but not with CD8+ T-cells. We additionally used LDA as a classical approach in pattern recognition to define the cell subsets that contribute most to favorable outcome during the recovery phase [23]. A high value in AUC of CD4+ T-cells and NK-cells after RT end was found to correlate with a PFS≥12 mo. High AUC of CD4+ T-cells and NK-cells as well as addition of ICI contributed mostly to favorable PFS, and a trend of fewer local recurrences in these patients was detected.

We also analyzed IL-6 plasma concentrations during and after treatment. In in-vitro experiments using human NSCLC cancer cell lines as well as mouse models, Yamaji et al. [24] found an association between IL-6 and tumor proliferation. The pro-inflammatory cytokine IL-6 has predictive potential of severity in a variety of diseases, e.g. heart disease [25], pneumonia [26]. Absolute IL-6 values are challenging to compare across individuals because of interindividual differences as well as dependency on gender, sex, and age [27]. Therefore, we interrogated the intra-individual dynamics of IL-6 within a 6-mo period following RT/cCRT using the standard deviation as a measure for the magnitude of this variation. We found a strong negative correlation of IL-6 variation with PFS, which is consistent with the findings of Yamaji et al. [24].

The current study has limitations. The study encompassed only a small number of patients (n=20) distributed over different treatment regimes. LDA may be problematic when used with small sample numbers [28]. To mitigate this issue, we interrogated a potential association with clinical benefit (OS, PFS) by various means including comparisons of single markers at individual timepoints or time intervals, as well as multi-parametric comparisons using unsupervised clustering and discriminatory analysis. Despite its limitations, our study has its value as it was conducted prospectively, and samples were collected longitudinally at multiple timepoints to characterize the recovery phase after RT. The results can be considered as a hypothesis to be tested in follow-up studies. If confirmed, the two identified parameters, peripheral CD8+ T-cell counts and IL-6 plasma concentration after treatment, are immediately amenable to clinical practice. A follow-up study is in progress including 40 inoperable stage III NSCLC patients [29].

## Conclusions

The present findings suggest that two parameters, which are commonly assessed in clinical routine, can be used to predict patient outcome. An early increase in CD8+ T cell lymphocyte count and low standard deviation in IL-6 plasma concentration are significantly correlated to patients with favorable, respectively poor outcome (PFS) after definitive therapy independent of treatment scheme. If confirmed, the two identified parameters are immediately amennable to clinical practice.

## Declarations

### Ethics approval and consent to participate

The study was approved by the Human Ethics Committee of the Ludwig-Maximilians-University of Munich reference no. 17-632 and conducted in accordance with the Declaration of Helsinki. Patients were included into the study after informed consent.

## Supporting information

NSCLC Supplemental Data

## Data Availability

All data produced in the present study are available upon reasonable request to the authors.

## Abbrevations

AB: Antibody
ALC: Absolute Lymphocyte Count
ASP: Antibody Staining Panel
cCRT: concurrent Chemoradiotherapy
Durva: Durvalumab
FCM: Flow Cytometry
FCS: Fetal Calf Serum
FL: Fluorochrome
Gy: Gray
ICI: Immune Checkpoint Inhibition
IQR: Inter Quantile Range
LDA: Linear Discriminant Analysis
LQR: Lower Quartile Range
mo: Month(s)
Nivo: Nivolumab
NLR: Neutrophil-Lymphocyte-Ratio
NSCLC: Non-small cell lung cancer
OS: Overall Survival
PFS: Progression-free Survival
RT: Radiation Therapy
TRT: Thoracic Radiotherapy
UQR: Upper Quantile Range
y: Year(s)

## Acknowledgements

We thank patients and their families, the clinicians and staff member of the LMU hospital, and colleagues at the Asklepios Fachkliniken München-Gauting.

For excellent support, we thank Anna Herbstritt, Barbara Mosetter, and Adam Slusarski.

