## Supplementary material for "Early recovery of leukocyte subsets is associated with progression-free survival in patients with inoperable stage III NSCLC after multimodal treatment: a prospective explorative study": NSCLC Supplemental Data

Supplement Text, Table Legend, and Figures Legends

Table S1

Tab. S1 Specifications of antibodies used for identification of leukocyte subpopulations

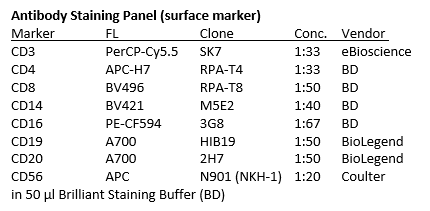

Table S2

**Tab. S2** Medians of absolute cell counts (Gcells / L) of leukocytes at time points RTend, C.1, C.2, and C.3 within the favorable PFS group (PFS ≥12 mo). Median; RTend n=8, C.1, C.2, C.3 n=9; Student’s t-test, * p<0.05, ** p<0.01

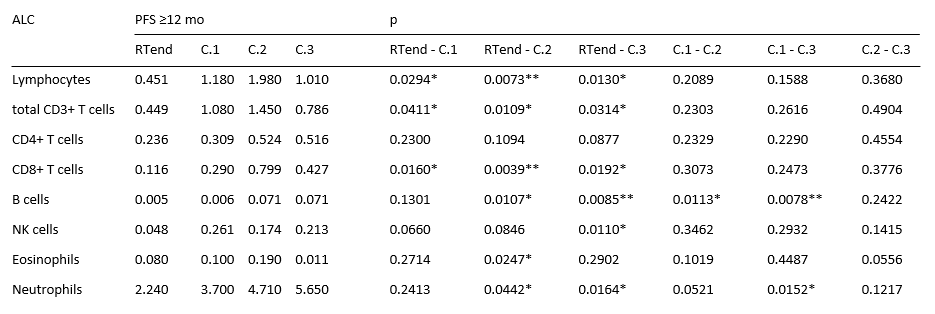

**Figure S1**

**Patient characteristics, study design, data collection**

20 patients were included in this trial. Two patients received RT alone, 18 patients were treated with platinum-based concurrent cCRT, and 7 patients received additional ICI either concurrently (nivolumab, 480 mg every two weeks, induction therapy with nivolumab prior to cCRT) or sequentially (durvalumab, 10 mg per kg body weight every 2 weeks for up to 1 y) to cCRT. In all patients, RT was applied with a median cumulative dose in equivalent 2 Gy fractions (EQD2) of 64 Gy (range 52-65 Gy). Two patients treated with cCRT received pembrolizumab salvage therapy after time points C.1 and C.2, respectively. Thus, subsequent time points of these patients were excluded from the analysis. One RT alone patient received a tyrosine kinase inhibitor between C.4 and C.5 for 8 weeks.

During treatment, 6 patients developed distant metastases (DM) while 9 had local recurrences (LR). Seven patients died during follow-up (within 1 y after the end of RT), 18 patients survived at least 6 months (mo), 14 patients at least 12 mo.

Blood was drawn on different time points before, during and after treatment up to 1y follow up, indicated below the timeline. For seven patients, blood of all time points could be collected. Others either missed a time point, revoked consent, or suffered from fatal disease progression. Data from time points after receiving salvage therapy using pembrolizumab of two patients were excluded from analysis examining impact of ICI-treatment.

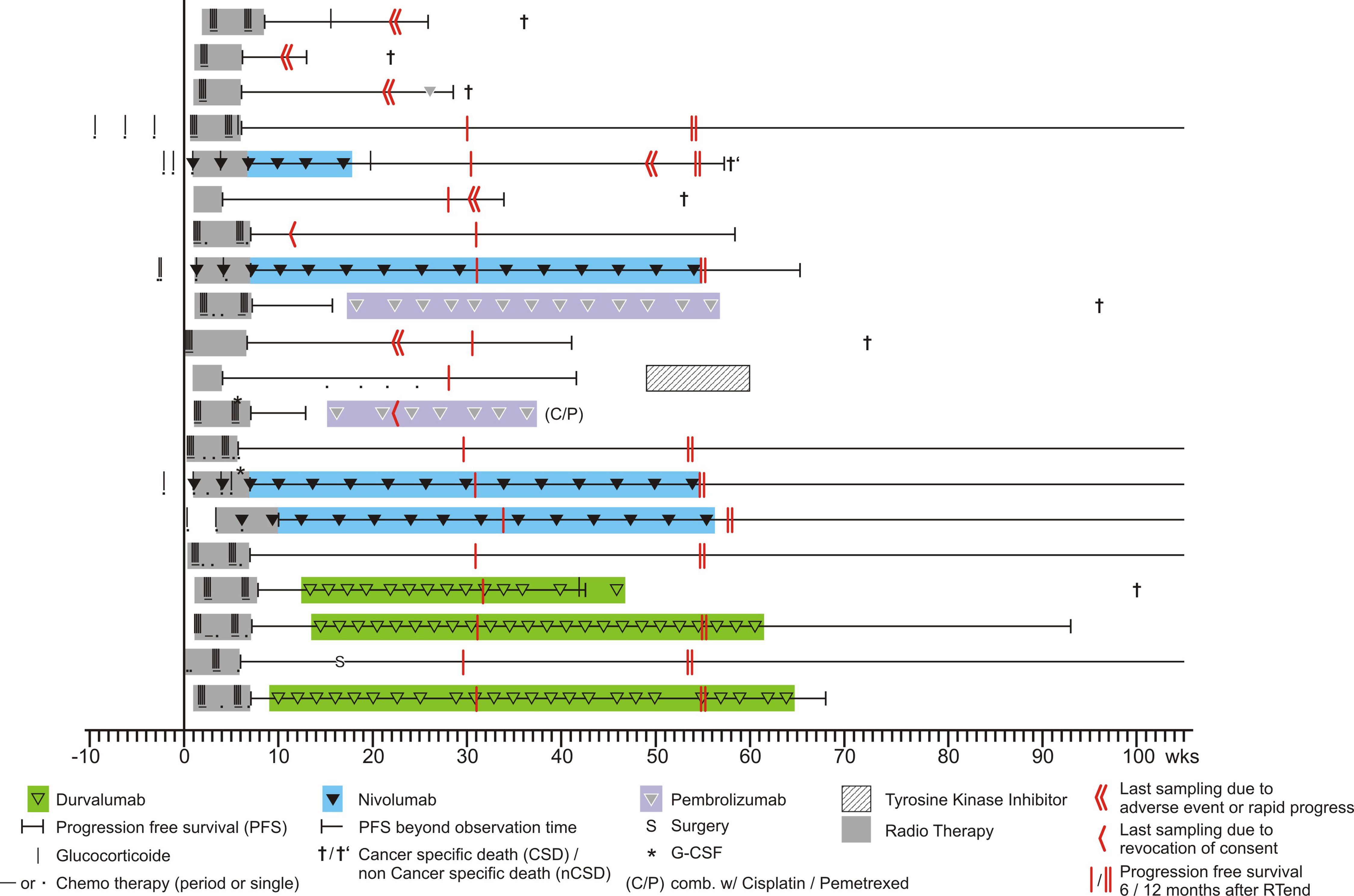

**Fig. S1** Summary view showing 20 patients (in lines), the timeline of treatment, medication, progression free survival, and overall survival (horizontally to the right).

Figure S2

**Sample Preparation and Gating Strategy**

Blood was collected using 3K-EDTA S-Monovette tubes (Sarstedt, Germany) and samples were analyzed within 3 hours. Using 100 µl of whole blood, erythrocytes were lysed using Q-Prep Workstation (Coulter, Germany). Cells were washed in 1 ml and resuspended in 100 µl FCM-Buffer (PBS without Ca^2+^ and Mg^2+^, supplemented with 2 mM EDTA and 2% FCS). For immuno-phenotyping, blood cells were stained on ice in the dark with a surface marker antibody panel to assess main blood cell subpopulations (Tab. S1). Cells were suspended in 250 µl FCM-Buffer, 50 µl (half the volume of blood) FlowCount fluorospheres (Beckman Coulter) were added for absolute cell count determination and analyzed on a Beckton Dickinson LSR II flow cytometer with Diva Software (version 8). A compensation matrix was generated using BD CompBeads, for flow data analysis FlowJo Software (version 10) was used. The staining protocols were established prior to the experiments, the formula for calculating the absolute cell numbers was (specified count of FlowCount beads (/µl) * cell count in assay) / (count of FlowCount beads in assay * 2).

To identify immunocyte subpopulations, leukocytes were gated within FSC-A vs. SSC-A to exclude debris. T-cells were defined as CD3+CD14-CD16-CD19/20- singlets (FSC-A vs. FSC-H) and subsequently subdivided into CD4+ and CD8+ T-cell populations (Fig. S2). B-cells were identified as CD19/20+ CD14- CD56- CD16- singlets. For NK-cells, highly CD16+, CD3+, CD19+20+ and CD14+ cells were excluded, and the remaining singlets cell population was gated for CD56+ CD16+. FlowCount (Beckman Coulter) counting beads were gated within FSC-A vs. SSC-A and subsequent event count was determined within 695nm emission channel (PerCP-Cy5.5) vs. SSC-H dot plot.

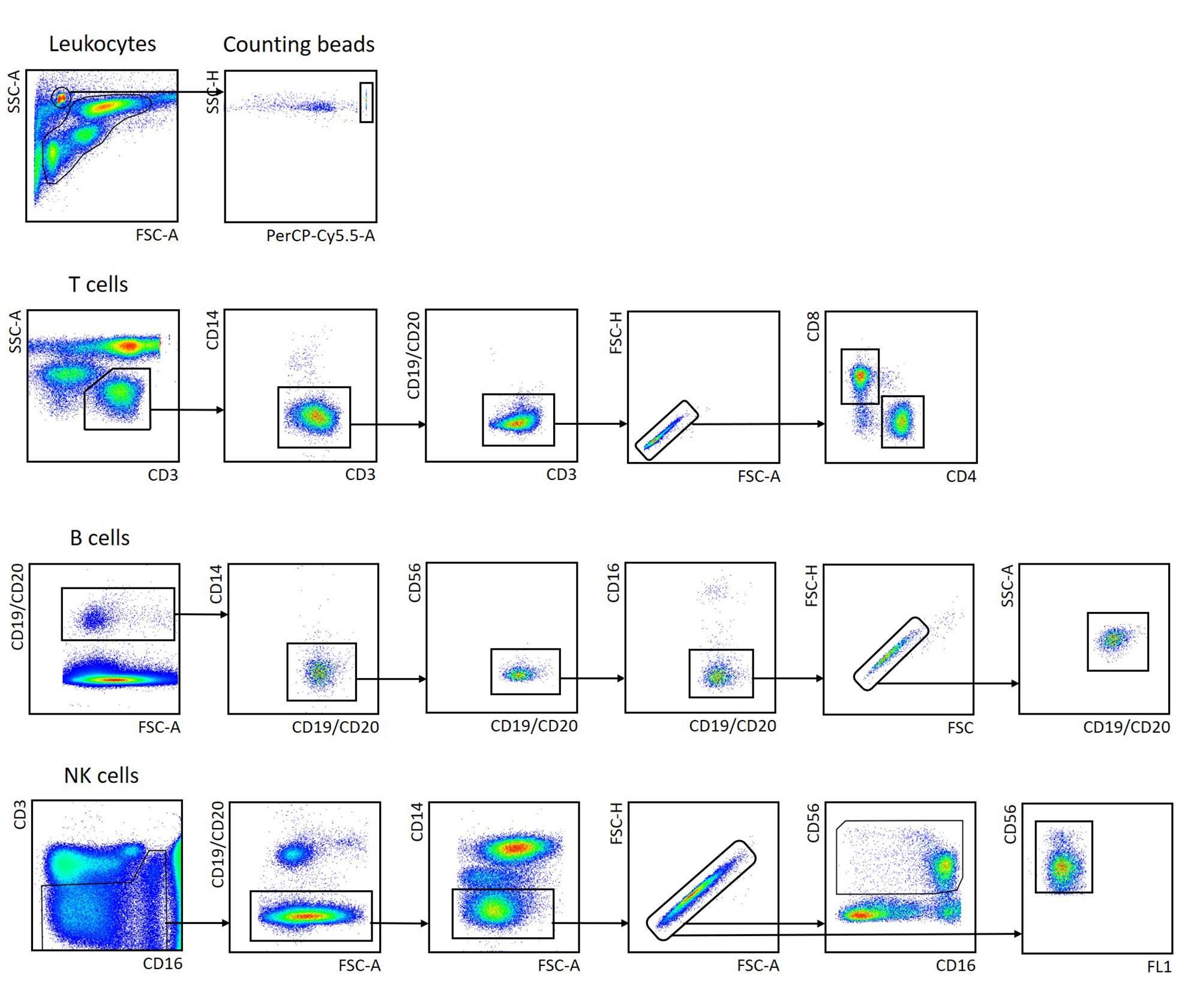

**Fig. S2** Gating strategy for the determination of the different cell population in peripheral venous blood. In each run counting beads were included as internal standard to calculate absolute cell counts.

**Figure S3**

**PFS & Overall Survival of patients with high, medium and low (UQR, IQR, LQR) lymphocyte cell counts at time point A.1 or C.3 as well as treatment groups**

Absolute lymphocyte counts (ALC) at A.1 trend to predict OS (b) but not PFS (a) (positive correlation). At time point C.3 (6 mo post-RT/cCRT) higher ALC trend to predict a longer OS (d). Looking at treatment groups (e, f) addition of ICI is beneficial for PFS and OS.

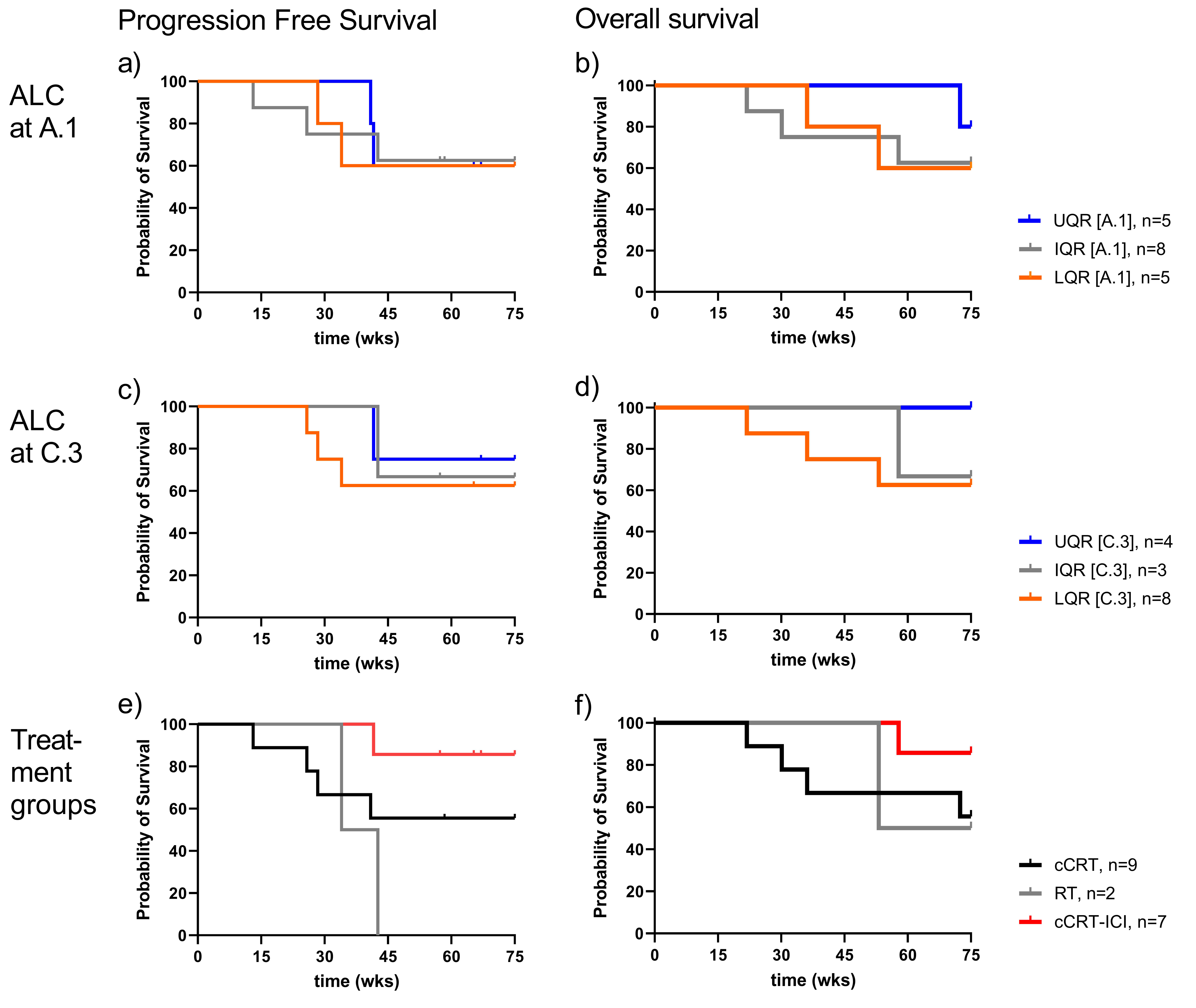

**Fig. S3** Survival Analysis (PFS, OS) of patients grouped according to the LQR, IQR, and UPR of the ALC at time point A.1 (a,b), and C.3 (c,d) as well as treatment group (e,f). For statistical analysis Logrank test for trend was performed, a) p=0.989, b) p=0.467, c), p=0.565, d) p=0.185, e) p=0.199, f) p=0.199. Numbers of total patients at risk see legend.

**Figure S4**

**Dynamics of leukocyte subpopulations after RT**

Exploring the predictive value of absolute count (AC) in lymphocyte subpopulations at the time point RTend and time points during follow-up revealed that the favorable PFS group (≥12 mo) showed significant increase in AC from RTend to C.2 (3 mo after treatment end) for B cells, eosinophils, and neutrophils. From RTend to C.3 (6 mo after treatment end) B cells, NK cells and neutrophils were significant (p>0.05).

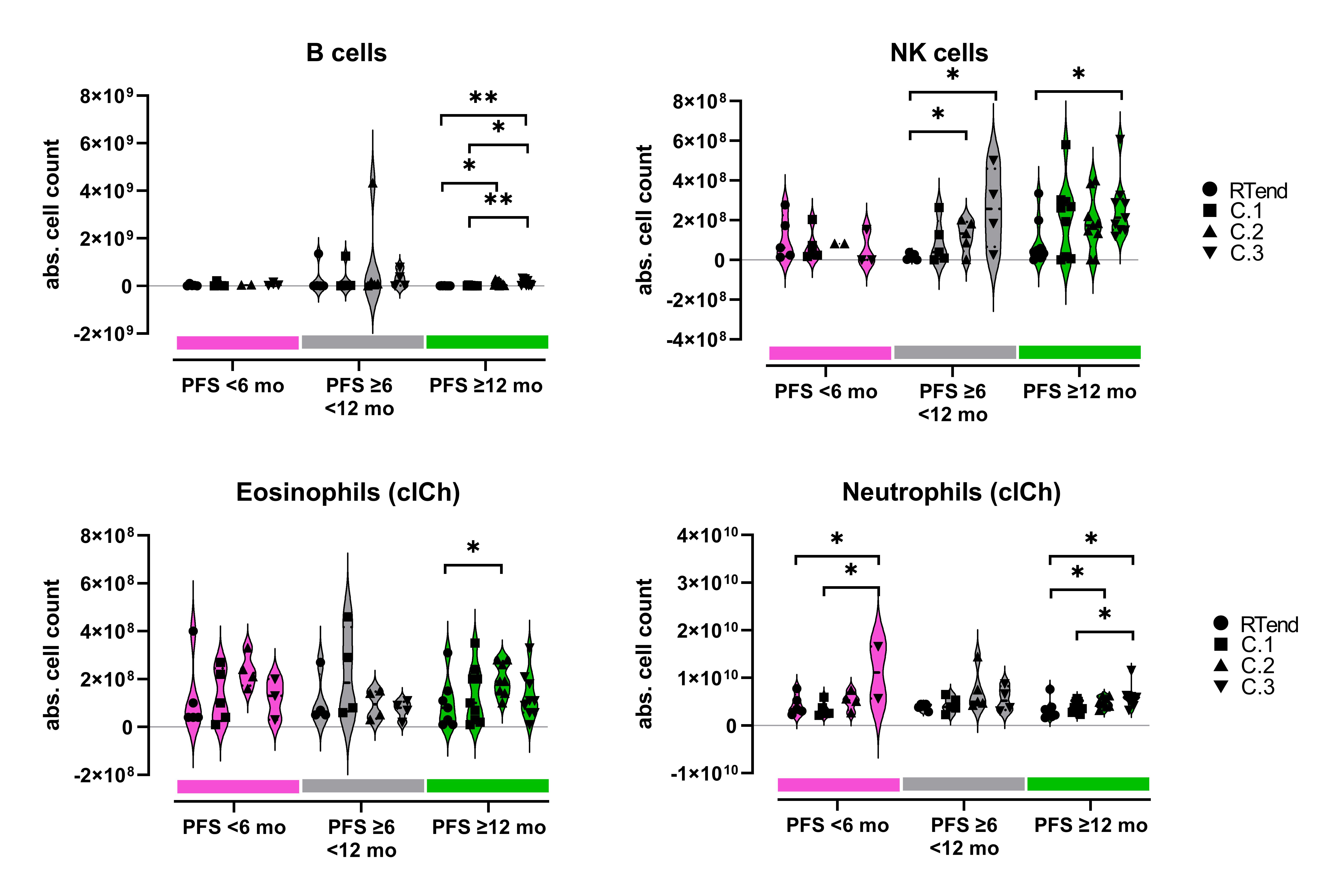

**Fig. S4** Absolute counts of lymphocytes and lymphocyte subpopulations at the time points RTend, C.1, C.2, and C.3 for the PFS groups (red PFS<6 mo, gray PFS≥6<12 mo, green ≥12 mo). Each dot represents an individual. Comparison was made between the abs. cell counts at the different time points within each PFS group. Cell counts were derived from flow cytometry (gating strategy in Fig. S2), neutrophil and eosinophil counts were determined by clinical chemistry routine lab (clCh). Bold dashed line in the violins indicate the median, the light dotted lines the lower and upper quartile. Student’s t test, * p<0.05, ** p<0.01.

**Figure S5**

**LDA using blood cell subpopulations separates patients according to PFS**

Partition plots of all predictor variable combinations are shown for a more detailed information on their discriminant contribution towards the PFS groups.

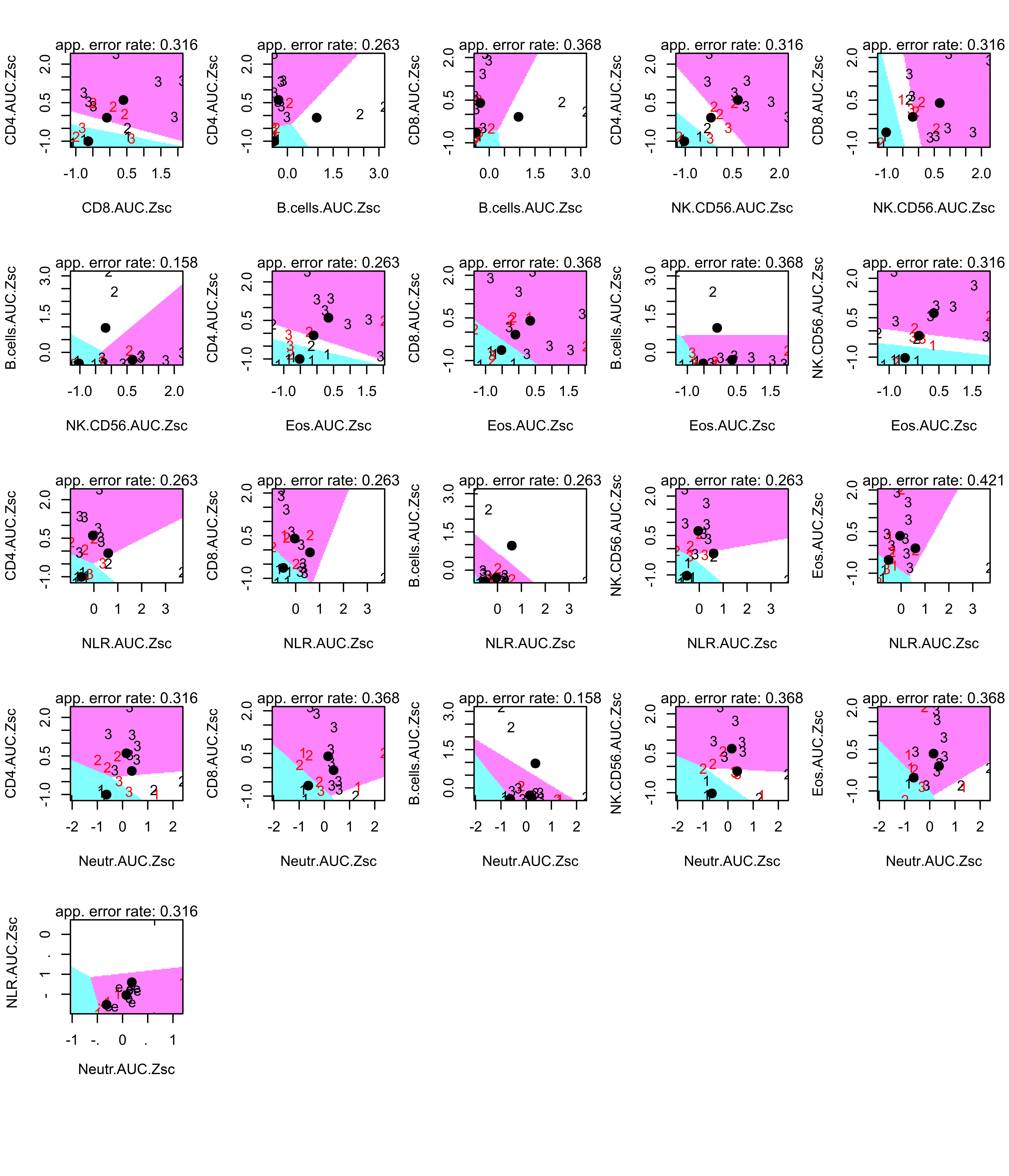

**Fig. S5** Partition plot of linear discriminant analysis including discrimination error rates of the different cell subtype combinations. AUC of cell counts of the interval from RTend to C.3 (6 mo after RT) were used and Z-score normalized. 1 represents PFS < 6 mo, 2 represents PFS≥6<12 mo, and 3 represents PFS≥12mo.
